# Restoring Cortically Mediated Movement and Sensation in Complete Tetraplegia

**DOI:** 10.1101/2025.08.19.25330198

**Authors:** Santosh Chandrasekaran, Sarah K. Wandelt, Aniket Jangam, Zeev Elias, Erona Ibroci, Christina Maffei, Isabelle A. Rosenthal, Richard Ramdeo, Joo-won Kim, Junqian Xu, Matthew F. Glasser, Allison Neuwirth, Todd A. Goldstein, Nathan E. Crone, Matthew S. Fifer, Gelana Tostaeva, Stephan Bickel, Douglas Griffin, Michael Funaro, Nicholas G. Carras, Rachel Pruitt, Netanel Ben-Shalom, Adam B. Stein, Ashesh D. Mehta, Chad E. Bouton

## Abstract

Spinal cord injury (SCI) affects millions worldwide, with over half of all cases resulting in tetraplegia, where a complete injury can cause profound motor and sensory loss in all four limbs^1^. Here, we demonstrate an artificial ‘double neural bypass’ (DNB) that integrates a bidirectional intracortical brain-computer interface with targeted spinal and brain stimulation to promote restoration of upper limb function in severe, complete paralysis. This hybrid assistive-therapeutic approach restores both hand movement and tactile sensation simultaneously via cortical mediation while promoting significant persistent sensorimotor improvements. The DNB uses a stable nested neural decoding architecture with deep reinforcement learning for fine grasping, along with patterned brain microstimulation (‘cortical mirroring’) and spinal cord stimulation to promote real-time and long-term functional recovery. Using the DNB, our participant with chronic C4 sensory/C5 motor complete tetraplegia regained the ability to self-feed, grasp delicate objects, and experienced persistent recovery of arm flexion and wrist tactile sensation. These findings represent a major advance in restoring meaningful function after severe, complete SCI, demonstrating that bidirectional neuroprostheses combined with targeted brain and spinal neuromodulation can drive durable sensorimotor recovery and improve independence and quality of life.

## Main

Spinal cord injury (SCI) is a leading cause of paralysis with tetraplegia occurring in over half of all cases and regaining upper limb movement is consistently ranked by individuals with SCI as the highest priority^1,2^. Furthermore, complete injuries, where no voluntary motor or sensory function is present below the level of injury, are particularly difficult to treat and recovery of useful function is exceedingly rare^3^. Previously, we and others demonstrated that a motor neuroprosthesis using a unidirectional intracortical brain-computer interface (iBCI) linked to real-time muscle stimulation can restore functional hand movement in complete tetraplegia^4,5^. However, these approaches did not include tactile sensory feedback, important for skilled grasping, or methods to promote neuroplasticity and persistent recovery. Tactile feedback via intracortical microstimulation (ICMS) of primary somatosensory cortex (S1) has also been previously used with robotic and virtual systems, but not while a user is moving their own limb^6^. Furthermore, while temporary enhancement of tactile sensation using ICMS has been demonstrated, persistent recovery has remained elusive^7^. Lastly, iBCI-mediated spinal cord stimulation has been demonstrated to recover function, but it was restricted to the lower limb and in incomplete SCI^8^.

To address the unmet clinical need of restoring upper limb function in individuals with high level, complete spinal cord injuries, we developed a ‘double neural bypass’ (DNB) approach – a hybrid system that can provide both assistive and therapeutic benefits. To achieve this, the DNB integrates a bidirectional iBCI to record and stimulate primary sensorimotor cortex, a stable neural decoding architecture to infer user intentions, deep reinforcement learning to support precision movements, and transcutaneous spinal cord stimulation (tSCS) to modulate targeted dorsal spinal roots for promoting neuroplasticity and persistent recovery. The bidirectional iBCI supports both assistive and therapeutic modalities, while tSCS supports therapeutic modalities exclusively. In our first-in-human DNB clinical trial spanning more than 3 years (Extended Data Fig. 1), we enrolled a male participant with complete C4 sensory/C5 motor tetraplegia resulting from a diving accident that had occurred 13 months prior. With an inability to lift his arms or hands to his face, hold objects, or feel any sensation in his distal forearms and hands, his upper limb sensorimotor functions were severely limited.

### Overview of the double neural bypass

To restore upper limb functional ability in our study participant, the DNB has several key subsystems serving during assistive and/or therapeutic modalities. To restore cortically mediated high precision hand grasping (assistive mode), the DNB uses multiple neural networks (Fig. 1a) in a nested closed-loop control architecture that utilizes a long short-term memory (LSTM) neural network and a deep reinforcement learning (RL) agent. Here, movement intentions are decoded from neural activity recorded via two microelectrode arrays implanted in primary motor cortex (M1) while specific stimulation patterns are delivered to up to three microelectrode arrays implanted in S1 (Fig. 1b). Functional magnetic resonance imaging (fMRI) and cortical parcellation from the Human Connectome Project (HCP)^9^ was used to localize cortical regions of hand representation and plan implantation locations prior to surgery, with intraoperative cortical stimulation refining the target locations (Fig. 1b). Example M1 intracortical activity used as LSTM inputs for different hand movements are shown in Fig. 1c, and example tactile sensory percepts evoked by various S1 stimulation locations are shown in Fig. 1d. For both motor and sensory therapeutic modalities, tSCS is delivered to a custom electrode patch^10^ (Fig. 1a) and for sensory intervention, S1 recording and stimulation is performed through the bidirectional iBCI.

**Fig. 1:**
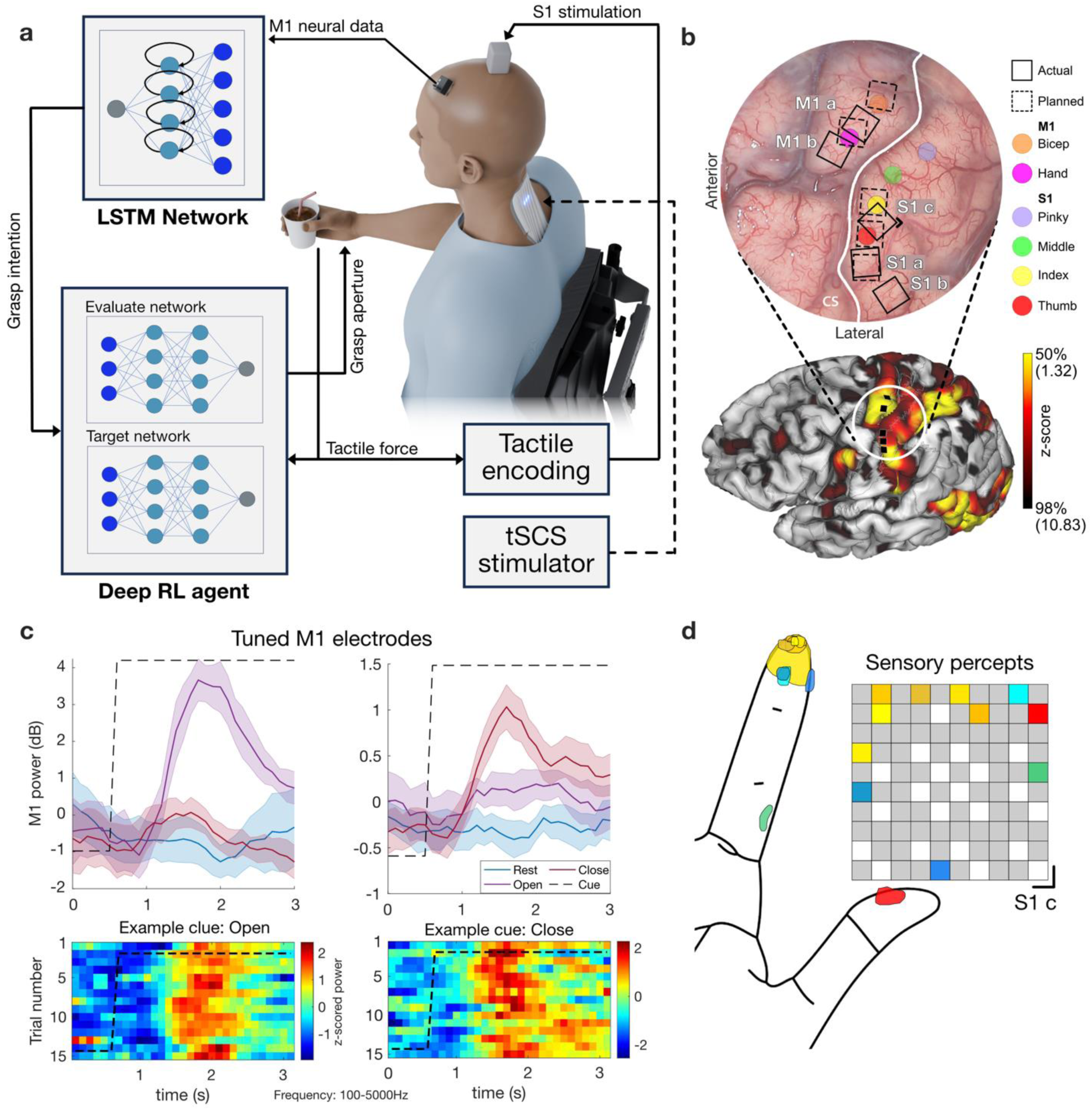
Double neural bypass system. **a)** Schematic showing the ‘Double Neural Bypass’ (DNB) system. **b)** Top: Intraoperative image of the brain showing M1 and S1 with central sulcus (CS) (white). Colored circles in M1 show intraoperative cortical stimulation sites that evoked EMG activation in right bicep brachii (orange) and twitches in the right hand (magenta). Colored circles in S1 show awake intraoperative cortical stimulation sites that evoked sensory percepts in thumb (red), index (yellow), middle (green), and pinky (purple) fingers. Dashed and solid squares show planned and actual locations for array implantation, respectively. The black notch in the corner marks the medial sensory array (S1c). Bottom: task fMRI response (z-score) to video-guided imagined and attempted thumb flexion and extension. Black squares are the planned array implantation sites shown on the pial surface using Connectome Workbench, overlaid on cortical areal boundaries from HCP cortical parcellation. **c)** Examples of M1 electrodes tuned to ‘Open’ hand (left) and ‘Close’ hand (right). Top: M1 power integrated in the 100-5000Hz range (n=15 trials, 95% CI). Bottom: z-scored power depicting each trial. **d)** Example S1-ICMS-elicited somatosensory percepts localized to the thumb and index fingers; corresponding stimulated electrodes in the depicted array are color-matched. The black notch on the bottom right corner marks the medial sensory array (S1c) as in panel B Top.

### Initial Spinal Cord Stimulation and Sensorimotor Baselining

For evaluating the efficacy of the hybrid DNB system, we first sought to establish sensorimotor baselines and assess the extent of functional recovery with tSCS alone in our participant. He presented with an inability to lift his arms and bring his hands to his face or generate volitional movement in the fingers, and this persisted during a 20-week baseline period, confirming he had plateaued in his recovery. It was also confirmed through sensory assessments using Semmes-Weinstein monofilaments (SWMs) that he lacked tactile sensation in both hands and wrists. We administered tSCS paired with activity-based training once a week by stimulating the dorsal cervical spinal column via a custom stimulator and an electronically controllable electrode patch^10^ (Fig. 2a) with anatomical landmarks ensuring consistent electrode placement (Fig. 2b). Recruitment profile mapping along the rostrocaudal axis showed that tSCS targeted at the C5-C6 spinal roots strongly recruited proximal as well as distal upper extremity muscles (Fig. 2c), but the distal muscles including the flexors (FDS) and extensors (EDC), were more selectively recruited when stimulation was targeted at the C7-C8 spinal roots (Fig. 2c).

**Fig. 2:**
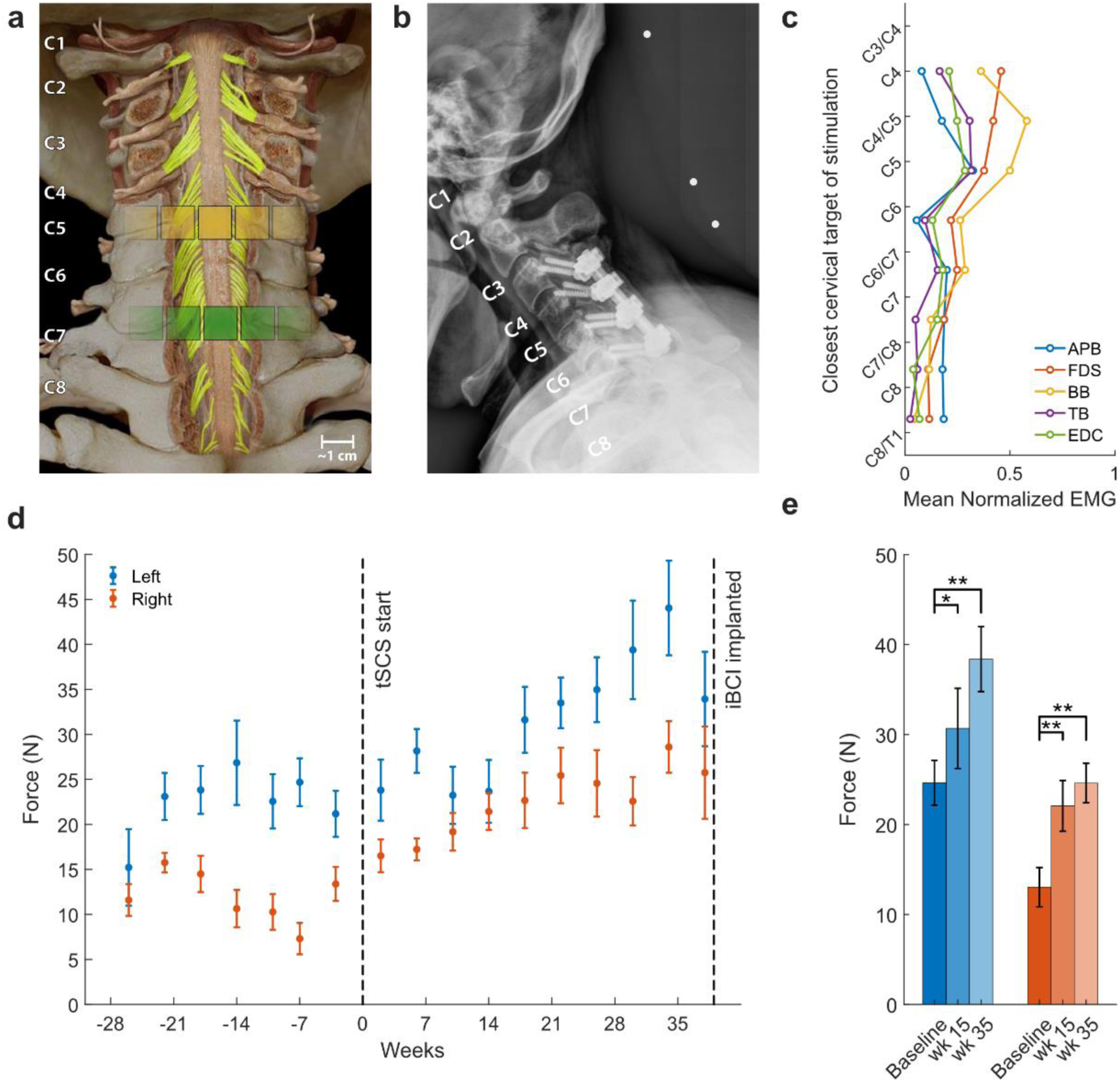
Persistent recovery of upper limb motor function by tSCS. **a)** Schematic showing the location on the cervical spinal cord dynamically targeted by the custom tSCS stimulator and electrode patch; C5-C6 (yellow electrodes) and C7-C8 (green electrodes) spinal root. **b)** Cervical spinal x-ray (note the surgical plate and screws for the C3-C6 spinal fusion) and the location of the electrodes. **c)** Recruitment profile of the muscles of the upper arm (APB=abductor pollicis brevis, FDS=flexor digitorum superficialis, BB=biceps brachii, TB=triceps brachii, EDC=extensor digitorum communis). **d)** Gains in volitional isometric elbow flexion force for left (blue) and right side (red). **e)** Mean gains in isometric elbow flexion force over 15- and 35-weeks post tSCS onset (*p < 0.05, **p < 0.01) for left (blue) and right side (red).

Based on the recruitment profile, tSCS was targeted at C5-C6 spinal root while performing isometric elbow flexion and extension. Similarly, tSCS was targeted at the C7-C8 spinal roots when the participant attempted to open and close his hands. Prior to initiating tSCS, assessment of volitional forces for 20 weeks established a stable baseline to preclude spontaneous recovery (Fig. 2d). We observed that, within 15 weeks of administering tSCS alone during activity-based training, there was a significant 70% and 25% increase in volitional force output (1-way repeated measures ANOVA with Tukey-Kramer HSD multiple comparison correction, p < 0.05) during isometric right and left elbow flexion, respectively. Further, over a period of approximately 35 weeks since initiating tSCS, the gains in volitional force reached up to 89% and 56%, respectively compared to the pre-tSCS volitional forces (same test, p < 0.01) (Fig. 2c, d). Moreover, this increase was persistent on both sides over several months and translated into a functional improvement wherein the participant could now bring both his hands up to his face.

However, the therapeutic tSCS targeted at the muscles of the hand and triceps did not result in any significant increase in strength or volitional control (Extended Data Fig. 2). Similarly, we did not observe any improvements to alleviate the tactile sensation deficits in the distal arm. These limitations of administering tSCS alone highlighted the need for a hybrid assistive-therapeutic solution to restore sensory and motor function to the most severely affected regions in complete SCI.

### Restoring cortically mediated hand movement and tactile sensation

We next shifted our focus to implementing the assistive portion of the DNB system to address the severely paralyzed hand muscles, and hand/wrist areas void of tactile sensation, that did not improve with tSCS alone. To restore cortically mediated grasping and tactile sensation simultaneously in the participant’s own hand, the DNB system combined real-time M1 decoding and tactile feedback through S1 microstimulation. Two intracortical M1 microelectrode arrays recorded neural activity (Fig. 1c) and, via NMES, drove the artificial activation of the muscles of the participant’s own hand. Force sensors placed in the palm of the hand provided somatotopically relevant tactile sensory feedback via S1-ICMS (Fig. 1a). Thus, we could provide motor control to, and sensory feedback from, the user’s own hand. The decoder for inferring grasp intent used M1 neural data during a task in which the participant attempted to open, close and rest his hand, guided by a virtual on-screen hand (Fig. 3a). The most consistent neural features determined by their mean correlation coefficient (MCC) served as inputs to a LSTM decoder (see Methods).

**Fig. 3:**
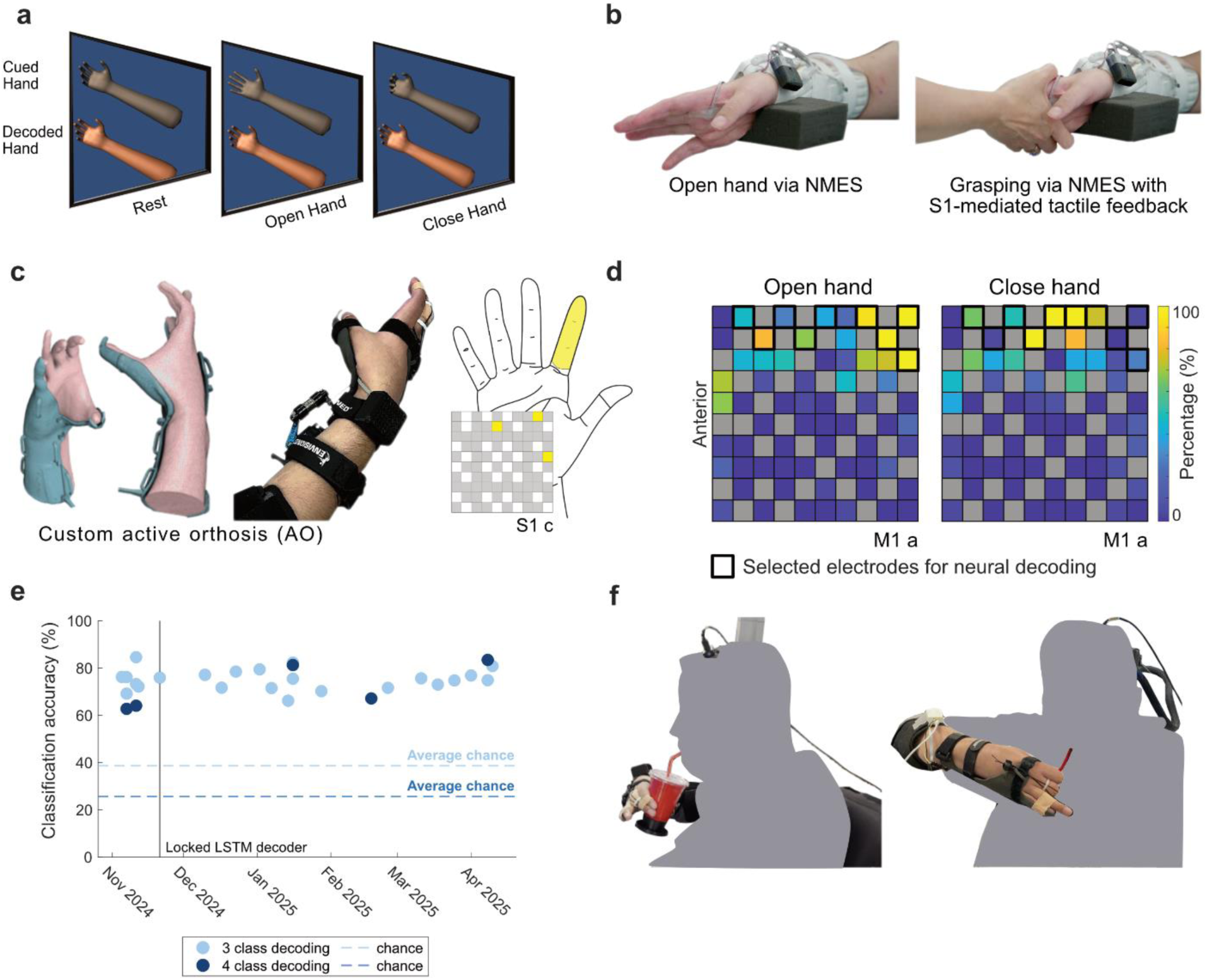
Closed-loop grasping with S1-mediated tactile feedback. **a)** Task design: Participant was instructed to imagine and attempt to rest, close or open his hand by following visual instructions on a screen (gray hand) and to hold the position for variable periods of time. **b)** Targeted activation of the participant’s own hand using cortically mediated NMES to perform open and close movements, in this case to grasp and feel another person’s hand. **c)** 3D printed low-profile active orthosis and the sensory percept activated during grasping. **d)** Percentage of sessions in which an electrode was tuned to Open vs. Rest (left) or Close vs. Rest (right) hand positions, out of 28 sessions (Kruskal-Wallis with post-hoc pairwise comparison Dunnett’s test, p < 0.05). **e)** A LSTM decoder was trained on 20 blocks of data and fixed over time using ten electrodes as features (see d, bold black border). Decoded conditions were rest, open, close (3-class) and reaching (4-class). Classification accuracies remained stable 5 months later. **f)** Activities of daily living such as drinking from a cup (left) and self-feeding (right) performed by the participant using both the gains obtained from tSCS therapy and the cortically mediated orthosis.

Electrode patches worn on volar and dorsal aspects of the right forearm provided NMES control of finger flexion and extension, respectively. The participant was able to cortically control the muscles of the hand using a LSTM-based decoder (Extended Data Fig. 3), allowing him to open (Fig. 3b, left) and close (Fig. 3b, right) his own physical hand. In one demonstration, he was able to grasp and feel his sister’s hand using his own, otherwise paralyzed and insensate hand (<u>Supplementary Video 1</u>).

While NMES was able to partially activate certain flexor muscles of the hand, others did not respond, which combined with nerve conduction study results suggested significant lower motor neuron dysfunction. To increase grasping forces, we developed a custom 3D-printed low-profile active orthosis (AO) that used an artificial tendon system to flex the index and middle fingers for naturalistic grasping. A force sensor embedded in the AO measured grasp strength, which was conveyed to the participant in real-time through S1 ICMS (Fig. 3c, see Methods). Decoded motor intent was used to activate the AO which modulated the grasp aperture. To further advance the DNB system for real-world use, we also developed a highly stable neural decoder. Training data was recorded on variable durations of opening, closing and resting his hand (3-class decoding). Additionally, a reaching condition, detecting when the participant was moving his arm, was added to mitigate unwanted decoder activations (4-class decoding). A preliminary decoder, trained on offline data of two sessions collected two days apart, provided feedback via a virtual hand (Fig. 3a) and was refined over four additional session days. As such, data recorded on 6 session days during a 16-day period helped train the stable decoder. Ten M1 electrodes, consistently active across trials (MCC ≥0.45 in ≥80% of training sessions), were selected as decoder features, and demonstrated significant tuning (Fig 3d, Kruskal-Wallis with post-hoc pairwise comparison Dunnett’s test, p < 0.05, left: open vs. rest, right: close vs. rest). The decoder demonstrated stable performance for all following experiments over a period of 5 months, achieving up to 84.6% accuracy (Fig. 3e). Finally, using the stable decoder and building on the motor gains obtained from the tSCS therapy alone, the cortically mediated AO enabled the participant to successfully perform activities of daily living (ADLs), such as drinking from a cup and self-feeding (Fig. 3f, <u>Supplementary Video 2</u>).

### High precision grasping with deep reinforcement learning

We observed that LSTM-based decoding of motor intent linked directly to AO control led to poor real-time grasp force regulation leading to under-or over-squeezing of objects. We also previously observed that M1 modulation is mostly phasic where strongest modulation occurs at initiation and termination of movement^11^. Inspired by the hierarchical nature of primate sensorimotor feedback loops^12^, we developed a nested neural network control approach to decouple grasp initiation and continued fine regulation (Fig. 4a). In this approach, the LSTM decoder infers grasping intention from M1 cortical activity patterns while a deep RL agent rapidly and continuously regulates grasping forces within specified limits. The participant was able to initiate grasping using the LSTM decoder, and the deep RL agent subsequently stabilized the grasp by modulating the grasp aperture based on the real-time tactile force received from the force sensor. The deep RL agent was trained in a simulated environment where the physical properties of the hand-object interactions were modeled numerically, and the deep RL agent was rewarded when forces were regulated tightly and responses were faster (see Methods). This novel RL-based framework supports adaptation and future learning as the user interacts with novel objects and refines grasping functionality.

**Fig. 4:**
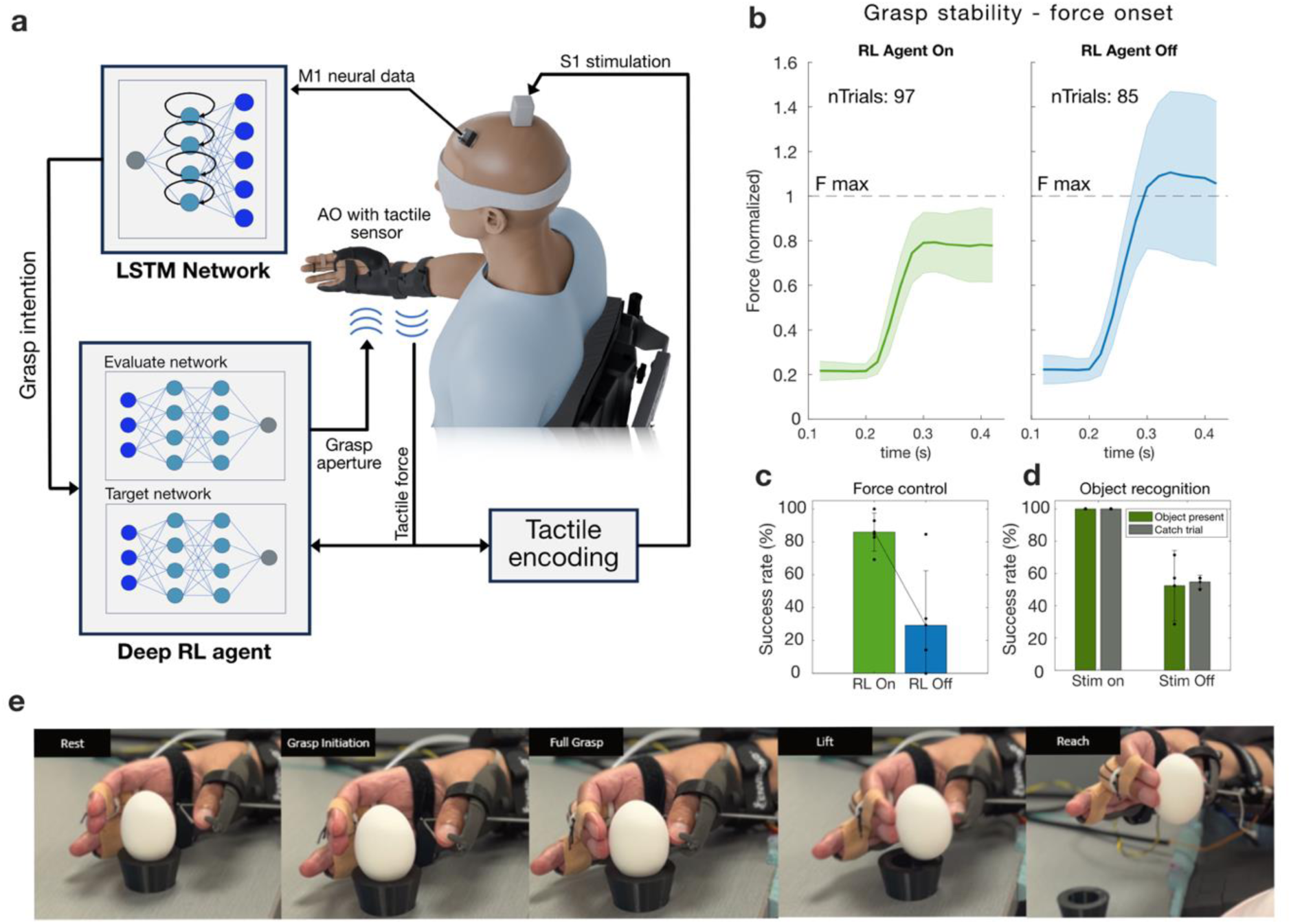
High Precision Grasping with Tactile Feedback. **a)** Experimental setup of the nested neural control architecture using reinforcement learning for closed loop grasping. **b)** Average force sensor data with standard deviation in the RL Agent on (left) and RL Agent off (right) conditions. Data were normalized to a predefined force limit. **c)** The participant achieved successful force control if the measured force remained under the instructed maximum force level, achieving 87% success in the RL Agent condition compared to 27% without it (unpaired t-test, p = 0.012). **d)** The participant achieved successful object presence recognition when he lifted the hollow eggshell only when he determined that an object was present. With S1-ICMS feedback, he achieved 100% success in both objects and catch trial conditions. Without S1-ICMS, he performed at chance level. **e)** Demonstration of precision grasp and lift of a hollow eggshell using the AO.

To test the nested LSTM-deep RL approach, we set up a task involving hollow eggshells. The participant was blinded to the eggshell and his hand but could see a real-time plot of the grasp force and was instructed to maintain it within a pre-defined limit; when the deep RL agent was employed, he was able to grasp the eggshell with consistent and safe force levels (Fig. 4b, left). Without the deep RL agent, the force generated was highly variable and often higher than the pre-defined limit (Fig. 4b, right). The deep RL agent allowed for a significantly higher number of successful trials compared to without it (87% success with RL, and 27% success without RL; p = 0.012, unpaired t-test), demonstrating higher successful force control (Fig. 4c, left). Lastly, the deep RL approach was used with and without S1-ICMS-based somatosensory feedback, to indicate if the eggshell was within the participant’s grasp. Visually blinded in both cases, the participant achieved a significantly higher success rate (100% vs chance level success without feedback, p < 0.01) when the S1-ICMS-based somatosensory feedback was employed, only lifting his arm when he felt he had the eggshell in his grasp (Fig. 4d).

### Persistent recovery of somatosensation

We next aimed to restore tactile sensation in the distal C6-C7 dermatomes, specifically the hand and wrist, since the previous 39-week tSCS regimen had not led to a significant change. Using the DNB system, we identified consistent spatiotemporal patterns of cortical modulation in contralateral S1 during imagined (and later, during physical tactile stimuli in the hand and wrist). We reasoned that repeatedly pairing this spatial activation of S1 with peripheral stimuli could promote short and/or long-term plasticity, potentially leading to persistent recovery of sensation. For this, we delivered stimulation to S1 with the same spatial pattern as the robust activity observed during imagined or physical touch. We termed this intervention ‘cortical mirroring’ (CM) since it essentially mirrored, or ‘played back,’ the natural spatial activation in S1. CM was applied after tSCS priming and this is referred to as the ‘tSCS+CM’ intervention described below.

During the tSCS+CM intervention, the participant engaged in motor and sensorimotor imagery tasks aimed at enhancing impaired sensorimotor processes (Fig. 5a). As the first step in the cortical mirroring approach, neural activity was recorded while the participant watched hand/wrist stroking and imagined corresponding sensations on four different locations on his right hand: index pad, index volar pad, thumb pad, and wrist. Next, we analyzed the integrated broadband power (100-5000 Hz) for each of the 96 electrodes in S1 and identified up to 16 electrodes (per hand/wrist location) that had the most consistently modulated power (highest MCC > 0.4). To raise spinal cord excitability, we applied tSCS over C5 or C7 spinal roots for up to an hour each, as was done previously (tSCS regimen) in the study. After briefly priming S1, which consisted of delivering mirrored patterns without task video or vibrotactile stimuli, we performed CM paired with vibrotactile stimulation for 5-10 minutes where we stimulated the electrodes previously identified. The index volar pad location on the hand served as a control, and no vibrotactile stimuli or CM/S1 stimulation was performed related to this location.

**Fig. 5:**
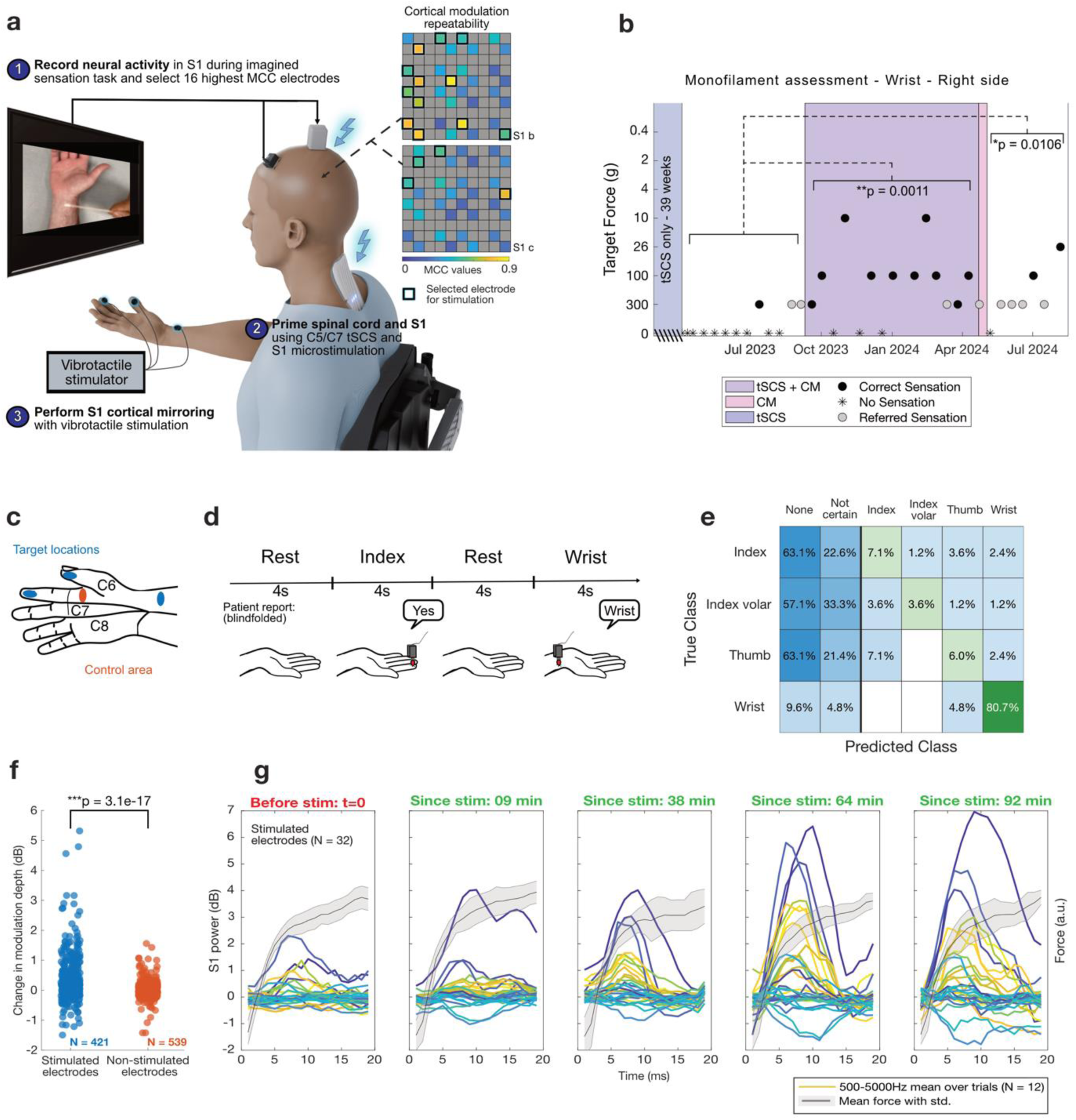
Persistent recovery of somatosensation. **a)** Experimental setup of sensory intervention. **b)** Sensory assessment results of the right radial wrist over time. Full dots represent the lowest target force the participant reported on a given assessment day, gray dots represent referred sensations, stars indicate no sensation. **c)** Target location for intervention: thumb pad, index pad, wrist pad, and index volar pad. **d)** Press and Hold experimental design. ∼700 g of force was applied to the blindfolded participant’s target locations, in random order, n=12. After each stimulus, the participant reported either the correct location, “No” if no sensation was perceived or “Yes” if touch was felt but the location was uncertain. **e)** Behavioral report confusion matrix over 11 session days. **f)** Change in depth of modulation before and after tSCS+CM intervention for stimulated and non-stimulated electrodes over 10 session days. Stimulated electrodes had a significant higher increase compared to non-stimulated electrodes (unpaired t-test, p < 0.001). **g)** Effect of time since stimulation on feature amplitude in stimmed electrodes on example session day (N = 32, one color per electrode). Neural data was aligned to force onset for each trial (mean and standard deviation of force in gray). Average electrode activity over all trials was plotted for 0.2 s before and 1.6 s after force onset. Maximum feature amplitude in stimulated electrodes was observed 92 min after first stimulation was applied.

We observed that tactile sensitivity, as assessed using SWMs on the radial aspect of the palmar side of the right wrist, significantly improved during the tSCS+CM intervention period (Fig. 5b). Here, the participant had significantly lower perception threshold of applied forces during the intervention period than during the baseline period (Wilcoxon rank sum test, p = 0.0011), perceiving as low as 10 g of applied force on the wrist. The post-intervention period presented more referred sensations (see Methods) and was significantly different from baseline as well (Wilcoxon rank sum test, p = 0.0106). The participant reported no sensation at the index pad, index volar pad, and thumb pad bilaterally across all sessions across all experimental phases.

These results suggest that the tSCS+CM intervention induced long-term plasticity changes in wrist sensitivity. To study the effect of the intervention on S1 modulation during tactile stimuli, a “Press and Hold” task was developed. Here, the blindfolded participant’s neural and verbal feedback responses were evaluated while 700 g of force was administered to the four previously described hand locations (Fig. 5c). The participant provided verbal feedback on whether he felt the sensation and if it matched the indicated location (Fig. 5d). The participant could feel his wrist with a high average accuracy of 81%, in contrast to other locations, over 11 sessions (Fig. 5e).

Cortical modulation corroborated the monofilament (Fig. 5b) and behavioral results (Fig. 5e), with significantly higher MCC values for touches compared to no touches (paired t-test, p < 0.001), but only for the wrist location (Extended Data Fig. 3a). Furthermore, stimulated S1 electrodes had significantly enhanced modulation to touch after the tSCS+CM intervention, in contrast to non-stimulated electrodes (Fig. 5f, unpaired t-test, p < 0.001). Importantly, the “Press and Hold” experiment, repeated at various intervals post-stimulation, showed feature modulation effects peaking 92 minutes after initial stimulation (Fig. 5g). Such modulations were absent in unstimulated electrodes (Extended Data Fig. 3b), suggesting short-term, electrode-specific, plasticity effects potentiated wrist sensitivity enhancement.

## Discussion

Paralysis can drastically impair independence and quality of life. For those living with a high-level SCI resulting in tetraplegia, regaining hand function is a top priority, above walking, bowel, bladder, or sexual function^13^. Furthermore, with a complete spinal cord injury, options for functional recovery are extremely limited. To address this important clinical need, we conceived and demonstrated an artificial double neural bypass that combines a bidirectional iBCI with targeted brain and spinal cord stimulation to restore hand movement and tactile sensation simultaneously while providing the benefit of persistent, functional recovery in specific areas as well.

Bidirectional iBCIs have made tremendous progress in decoding motor intent to control robotic or virtual arms and in restoring the sense of touch as these artificial arms interact with objects^6,14^. However, restoring cortically mediated sensorimotor function in the user’s own limb is unexplored. Furthermore, our work and that by others has shown that muscles lacking a minimal level of baseline force in individuals with a chronic complete SCI, typically do not exhibit functional recovery following spinal cord stimulation interventions^10,15–18^. We therefore developed a hybrid system that combines a bidirectional iBCI-mediated neural bypass SCS with both assistive and therapeutic modalities to address even chronic motor and sensory complete injuries.

With the DNB system, the participant was able to complete independent ADLs such as self-feeding and drinking, hold and feel a loved one’s hand, and perform delicate, high-precision grasping functions such as picking up hollow eggshells without damaging them.

Outside of study sessions, the long-term gains from the DNB system have enabled the participant to independently scratch and wipe his face. Notably, he has also reported being able to pet his dog and feel her fur on his previously insensate right wrist. This evaluation of the DNB system was limited to one participant, but it demonstrated the potential for highly impactful benefits of such a hybrid neuroprosthetic-neuromodulation system for those living with complete spinal cord injuries.

One of the challenges of restoring iBCI-mediated motor control in the human hand has been achieving precise grasp regulation for real-world tasks (e.g. handling fragile objects and foods, etc.). Using a novel deep reinforcement learning approach, we demonstrated a proof-of-concept system capable of rapidly and accurately stabilizing the grasp force enabling successful interactions with delicate objects. Inspired by the distributed neural network architecture in primates^19^, we developed a nested control architecture combining a highly stable recurrent neural network and deep reinforcement learning to allow the participant to perform precision grasping tasks with high success. The participant also could complete the task while conversing with people in the room simultaneously, suggesting reduced cognitive load and future applicability of this approach in real-world settings.

In addition to motor improvements, we show that combining tSCS with cortical mirroring using microstimulation in S1 can improve somatosensation persistently in an individual with a sensory complete SCI. The interventions enabled the participant, with impaired sensation in his right wrist, to detect significantly lower force levels. This gain in overall sensitivity persisted for at least two months even after cessation of stimulation. It has previously been shown that S1 electrodes that elicit sensation when stimulated^20^ also modulate imagined sensations^21^. We found that neural modulation of stimulated electrodes to physical touches increased after S1 microstimulation, consistent with spike-triggered microstimulation results in primates^22^. This electrode-specific increase of neural modulation suggests intervention-dependent cortical plasticity may have contributed to sensitivity improvement. Touch to the other hand locations did not cause S1 neural modulation before or after intervention and did not improve over time. This suggests a minimum level of residual signal transmission from the spinal cord to S1 may be necessary to allow intervention-dependent sensory improvements, as observed for motor rehabilitation^18^. Additional refinement of tSCS and S1 stimulation patterns and their interaction may further improve sensory recovery using this new approach.

In this work, the assistive and therapeutic aspects of the DNB approach were used to restore functional and persistent motor abilities. Furthermore, a significant gain in sensory function was achieved when tSCS was combined with a cortical mirroring method. By restoring motor and sensory abilities more durably, the therapeutic approach can provide continued benefits to the user even when not actively using the system.

Restoring movement and sensation in the participant’s own hand allowed him to achieve ADLs and increased independence, while enabling socially and emotionally rewarding engagements such as holding and feeling the hand of a loved one.

Taken together, this work shows that combining a bidirectional iBCI with brain and spinal neuromodulatory approaches targeted at different points along the motor and sensory axis can enable functional and precision grasping tasks, while potentially unlocking persistent recovery of motor and sensory functions in the upper limb.

## Methods

### Study design and participant

The study is registered at ClinicalTrials.gov (NCT03680872) and was conducted under an Investigational Device Exemption (IDE G170200) issued by the US Food and Drug Administration (FDA). The protocol was approved by the Northwell Health Institutional Review Board (IRB# 17-0840) and was in accordance with the Declaration of Helsinki. The participant provided written informed consent for study participation, and permission for the use of photos, along with audio and video recordings, in which he appears.

The participant (male, in their 40’s at study enrollment) sustained a bilateral C4 sensory/C5 motor ASIA Impairment Scale A spinal cord injury following a diving accident and underwent a C3-C6 spinal fusion. At time of enrollment, the participant was 13 months post-injury, had undergone physiotherapy and required complete assistance with all activities of daily living. The study involved different phases as shown in Extended Data Fig. 1. For the first year of the study, the participant underwent activity-based training paired with transcutaneous spinal cord stimulation (tSCS) once a week. Surgical planning was performed using a 3T MRI scanner, followed by implantation of a brain-computer interface (BCI) and the study participant attended experimental sessions 1 to 3 times a week.

### Upper extremity clinical assessments

Upper extremity strength and sensation were assessed every two weeks. Strength was assessed using the motor examination for upper extremities protocol established by the International Standards for Neurological Classification of Spinal Cord Injury (ISNCSCI). Tactile sensation was evaluated using Semmes-Weinstein monofilaments (North Coast Medical, Inc.) designated as 6.65f, 6.10f, 5.46f, 5.07f, 4.56f, 4.31f, 3.61f, corresponding to the logarithm of applied force of 300, 100, 26, 10, 4, 2, 0.40 grams, respectively. Participant, while blindfolded, reported the perceived location of three 1.5-second taps per filament applied in descending order of filaments to randomized hand/wrist locations. “Correct” indicated accurate localization, “No Sensation” indicated absent perception, while “Referred Sensation” indicated a mislocalized perception. Starting with the 6.65f filament, testing stopped at the first absent or referred perception. Scoring was based on the thinnest perceived filament, with no sensation = 0, referred sensation = 0.5, 6.65f = 1, increasing by 1 for each filament (6.10f = 2, 5.46f = 3,…, 3.61f = 7)^23^. Changes in perceived sensations over experimental time periods were assessed using Wilcoxon rank sum tests.

### Activity based training - elbow extension and flexion

The general activity-based training protocol has been previously published^10^ and is briefly summarized here. The participant’s arm was placed on a custom-built board fixed with a tension/compression load cell (25 lb, Model M31, Honeywell International, Inc., USA), with the wrist strapped over the load cell to allow for isometric force measurement of elbow flexion and extension. The arm was extended forward and elbow flexed at 10° and 15° for flexion and extension tasks, respectively. Electromyography (EMG) activity and force were recorded for each task through Noraxon MyoResearch software (Noraxon USA, Inc) for abductor policis brevis (APB), biceps brachii (BB), triceps brachii (TB), flexor digitorum superficialis (FDS), and extensor digitorum communis (EDC) muscles. Each task consisted of two sets of five 5-second repetitions (flexion or extension) followed by 5 seconds of rest, with a 3 to 4-minute inter-set rest. Audio cues signaled the start and end of each repetition. The participant was verbally encouraged and provided a visual force goal to maximize and sustain force generation. All tasks were performed for both arms.

### Transcutaneous spinal cord stimulation (tSCS)

Transcutaneous spinal cord stimulation (tSCS) was delivered using a custom stimulator and an electronically-steerable electrode patch^10^. The stimulator consisted of an ESP32 feather microcontroller (Espressif Systems, China) that digitally generated a biphasic 10 kHz, 0.5 ms long (5 cycles), sinusoidal waveform repeated at 50 Hz. A voltage-driven digital input class-D audio amplifier (TAS5825PEVM, Texas Instruments) scaled the waveform to the desired stimulation amplitude. A 10:1 step-up transformer (42TM003-RC, Xicon) helped increase voltage output, provided signal isolation and impedance matching. Current limit circuitry was employed to ensure that the power density never exceeded 0.25 W/cm^2^. The stimulator was powered by a 24 V battery.

Consistent placement of the custom transcutaneous patch was ensured using the inion of the external occipital protuberance as a landmark. Two interconnected 5×10 cm hydrogel electrodes (Axelgaard Manufacturing Co.) placed along the lumbar spine midline served as return electrodes (anodes). To target specific cervical segments of the spinal cord, stimulation was provided simultaneously to 3 adjacent electrodes (electrode dimension: 10 mm x 10 mm, 1 mm pitch) within a single row, spanning the cervical midline.

### tSCS-mediated muscle recruitment

Recruitment profiles were generated by administering tSCS at 3 Hz. Increasing stimulation amplitudes (100-225 mA, in ∼25 mA increments) were applied to each of eight electrode rows for up to 30s while recording EMG. Snippets (5-55 ms post-stimulus artifact) with peak-to-peak amplitudes exceeding 5x baseline standard deviation were analyzed for each channel. For each muscle, peak-to-peak amplitudes were normalized to the maximum observed across all amplitudes and electrode rows. An X-ray image in the sagittal plane, with radio-opaque markers placed at 7 cm and 9.2 cm caudal to the participant’s occipital protuberance was used to determine electrode location relative to spinal roots and vertebral landmarks.

### tSCS with activity-based training

The experimental protocol began with 10 minutes of tSCS priming over the C5-C6 spinal root, during which the participant was asked to rest and not move during this time. This was followed by pseudorandomized left or right elbow flexion and extension tasks (as described above) with tSCS actively stimulating. Following this, 10 minutes of tSCS priming over the C7-C8 spinal root, followed by a bilateral hand open and close task wherein the participant is attempting to open and close his hands guided by a virtual hand, with the tSCS actively stimulating.

### Functional magnetic resonance imaging

The participant underwent scanning in a 3T MRI scanner (Prisma, Siemens, Germany) and a 64-channel head-neck coil with Human Connectome Project (HCP)-style^24^ structural and functional MRI protocols (for scanning parameters, see Supplementary Table 1). To localize hand representations in the sensorimotor cortex, the participant performed motor- and sensory-specific tasks during fMRI acquisition. While for M1, the task consisted of video-guided imagining and attempting of finger flexion (thumb, index, or middle), for S1, the task involved imagining sensation while watching videos of a Q-tip stroking the same fingers. Each task consisted of 12 seconds of activity (flexion or stroking), followed by 12 seconds of rest, with 10 repetitions. Additionally, for S1 tasks, a TENS unit (LG TecElite) delivered electrical stimulation (up to 10 mA, 30 Hz, 300 µs pulse width) to the same fingers using ring electrodes (Natus Medical Inc.) placed across the first knuckle. A programmed microcontroller activated solid state relays to target the correct finger and control the stimulation timing (12s ON, 12s OFF) to synchronize with the Q-tip videos. The sensory tasks were performed in 3 varieties: Q-tip video only, TENS only, and both simultaneously. However, the activation observed in S1 was weak and was ultimately not used for array implantation planning.

MRI preprocessing used the HCP minimal preprocessing pipelines (v3.27^24^) including motion and distortion correction, cortical surface reconstruction and subcortical segmentation, T1w/T2w-based myelin content and cortical thickness maps, fMRI data transformation to MNI and CIFTI gray-ordinate standard spaces using MSMAll-based registration^25,26^, and 6 mm surface smoothing. Spatially specific structured noise was removed using the HCP’s multi-run (v4.0) ICA-FIX^27–29^ with training data for multi fMRI (multiple finger tasks) and linear trends with a high pass filter (>0.1 Hz) without regressing out motion parameters. Somatotopic functional responses were estimated using a generalized linear model (GLM)-based fMRI analysis^30^ on the gray-ordinate data space for each finger.

Task fMRI activation maps, visualized in Connectome Workbench^9^, guided array implantation. Planned locations were identified as cortical vertices, with 4 mm borders, transformed into volume space ROIs (AC-PC aligned T1w) (*wb_command-surface-geodesic-rois, -metric-label-import,* followed by *-label-to-volume-mapping*). These volumetric ROIs were registered to the native T1-weighted image, thresholded to create binary masks and merged into a single label volume using *fslmaths*. The merged label was “burned” into the native T1 and exported as a *NIfTI* file (*nii.gz*) to ensure compatibility with the surgical navigation system (StealthStation, Medtronic, USA).

### Intraoperative cortical stimulation

Intraoperative motor and sensory mapping were performed via direct cortical surface stimulation using a 2 mm ball tip monopolar or bipolar probe, respectively. Under anesthesia, motor responses were elicited via 3 s long, 50 Hz, 4 mA, biphasic stimulation trains (pulse width: 100 μs), with high-impedance rejection enabled, and measured using EMG. Stimulation of superior and inferior primary motor cortex (M1) elicited EMG responses in the upper arm and frontalis muscles, respectively. Hand twitches in response to motor cortex stimulation were also noted and this location was marked for M1 array implantation. Furthermore, awake positive sensory mapping was performed to determine tactile percepts for various stimulation points in somatosensory cortex. Four of the fingers (Fig. 1b) were localized successfully and these responses were used to finalize the sites to target for S1 array implantation.

### Array implantation

The participant was implanted with five 10×10, 1.5 mm platinum microelectrode arrays with sputtered iridium oxide film (SIROF) coated tips (Neuroport Array, Blackrock Neurotech) in the hand areas of M1 (2×64-channel arrays, 128 total) and S1 (3 sparsely populated arrays, 32-channel each; 96 total) in the left hemisphere. Following M1 mapping under anesthesia and awake positive mapping of S1, the arrays were implanted using a high-speed pneumatic press. Both M1 arrays were connected to a single pedestal mounted anteriorly and all S1 arrays were connected to a second pedestal mounted posterior medial to the craniotomy, followed by a complex neuroplastics closure.

### Neural preprocessing pipeline

Broadband electrical activity was recorded from the NeuroPort arrays using Neural Signal Processors (Blackrock Microsystems). Analog signals were amplified and digitized at 10 kHz. Non-overlapping periodic Blackman windows of 1000 datapoints (100ms) were applied for frequency analysis, followed by a short-time Fourier transform (10 Hz resolution). Signal amplitudes were integrated across pre-defined frequency bands (100 to 5000 Hz), smoothed using a 200-millisecond boxcar filter, log compression for normalization and drift correction, standardized using a 24-second running mean and scaled to decibels (dB) by multiplying by 20. These frequency bands have been shown to exhibit highly repeatable amplitude modulation related to movement and tactile stimuli^11^.

### S1 mapping

S1-ICMS through SIROF microelectrode arrays using a Cerestim R96 microstimulator (Blackrock Neurotech) generated percepts at the fingertips of the participant. Sensory stimulation waveforms led with a cathodic phase of 100 µA, pulse width of 300 µs, and interphase width of 100 µs. Individual electrodes were stimulated at 50 Hz for 2 seconds to generate percepts in the palmar side of the index and thumb regions. The zone of perception was drawn on a hand image using a Python-based open source interface^31^, as verbally described by the participant.

### Standard virtual sensorimotor task - hand open/close

The participant performed an attempted movement task cued by a virtual hand (Unity Software Technologies) to open, close or rest his right hand. The cued followed a 4s cue (maintain movement), 4s delay (rest) paradigm. 15 repetitions of each motion (open or close) were performed. Cue orders were randomized.

### Decoder training

Decoder training data was collected over five sessions within a 16-day period using two paradigms.

Experiment 1 (18 data blocks): The participant mirrored virtual hand movements cued on a screen to open, close and rest his hand for variable durations (rest time: 1.5-11.5s, open hand: 1.5-4.5s, close hand: 1.5-5.5s).

Experiment 2 (4 data blocks): The participant performed cued hand closing, reaching up and down (emulating drinking from a cup) and opening his hand for variable durations (rest time: 4-13s, open hand: 3-6s, close hand: 3-6s, reaching: 5-6.5s).

The first 9 data blocks were recorded without feedback (offline). Subsequent 11 blocks included real-time decoding feedback of decoded hand movements in the form of a second hand on the screen. Long Short-Term Memory models were trained iteratively as more data became available.

Out of 22 blocks, 20 trained the final decoder, and two (one per experiment), tested the decoder. Only M1 data was used, as S1 was used for stimulation. Feature selection on training blocks used mean correlation coefficient (MCC) for each electrode. High MCC values indicate electrodes consistently and repeatably activated during each trial, whereas low MCC values suggest little correlation to the selected time-period of interest^11^. Trial data (0.2 to 1.6s after cue onset) for each electrode was normalized, and temporal correlation coefficients to the average of all trials were calculated, and averaged. 10 electrodes with an MCC ≥ 0.45 in ≥80% of training blocks were selected.

The long short-term memory (LSTM) network (200 hidden units, tanh activation, sigmoid gate, 0.01 dropout) predicted rest, open, close and reaching movement states every 100ms. Training used Adam optimizer (0.001 learning rate, 128 batch size, 200 epochs), categorical cross-entropy loss, and L2 regularization (0.0001 weight decay). This fixed model was used for all subsequent experiments (5 months period).

### Neural activity tuning analysis

Neural activity differences between hand conditions (rest, open, close) were assessed per electrode using the Kruskal-Wallis test. Significant differences (p < 0.05) prompted a post-hoc pairwise comparison Dunnett’s test between movement condition and rest and adjusted for multiple comparison using Bonferroni-Holm correction. An electrode was considered tuned for p < 0.05 for at least one movement.

### Closed-loop light grasping

Closed loop grasping was achieved through a hybrid system that combined long-lasting gross motor improvements with BCI mediated assistive technologies. An LSTM decoder (similar as described above, but trained only on one session day) was used to predict movement intentions. The decoded output was then wirelessly sent to a custom-built high-resolution neuromuscular electrical stimulator. The stimulator, secured at the wrist, delivered patterned stimulation (0.5 ms long, 10 kHz charge-balanced multiphasic pulses repeated at 50 Hz) through transcutaneous electrode arrays positioned over the muscles in the forearm responsible for finger flexion and extension. To prevent stimulation-based electrical artifacts from affecting the decoder, we created a hardware clock which would trigger the stimulation pulses for all devices and simultaneously send a trigger to the neural signal processor for 11.2 ms, generating a state to halt the data from being used for movement prediction until the amplifiers completely recovered. Tactile force feedback was implemented using Honeywell FMA 5 N force sensors affixed to the thumb and index tips, and the palm base. These sensors captured the pressure exerted on the hand, and transmitted the data to a computer, which used the incoming data as a trigger for sensory stimulation. This approach generated a sensation localized to the corresponding sensor location.

### Closed-loop precision grasping using deep reinforcement learning

A low-profile, custom active orthosis (AO) was developed to provide stronger grasping for object manipulation, while a passive elbow brace, to provide counter resistance, was worn. The nylon body of the AO was designed by 3D scanning (3D Shape, Copenhagen, Denmark) the participant’s hand and forearm, followed by the Spentys (Forest, Belgium) digital fabrication workflow to ensure anatomical conformity, proper grasp aperture, and comfort. Minor alterations were made in Autodesk Fusion (San Francisco, California) to account for the hardware mounted to the device. The AO was 3D printed (Formlabs Fuse 1, Sommerville, Massachusetts) using Nylon 12 Powder, selected for its high tensile strength and biocompatibility standards. A 14 kg servo motor (Injora, Guangdong Province, China) was mounted on the AO in the middle of the flexor digitorum superficialis region. The servo was coupled with a magnetic release to a cord which was attached to a 3D printed custom acrylic ring (Formlabs Form 2) using BioMed Clear Resin. The ring was worn on the middle finger, which was tethered to the index finger, enabling coordinated flexion. The grasp force was measured by a tactile force sensor (Honeywell FMA 5N) embedded between the AO and back of the thumb IP joint, such that all grasp forces were applied against the opposing two tethered fingers. Force sensor data was streamed to MATLAB at 50 Hz and was used to drive real-time sensory feedback. Based on the S1 mapping, three electrodes were selected for AO sensory feedback, which reliably elicited a strong tactile percept on the palmer side of his index finger. When grasp forces exceeded an object specific threshold for 200 ms, ICMS was delivered to these S1 electrodes at 100 µA (pulse width of 300 µs, interphase 100 µs, frequency 100 Hz) for 1-2s. To avoid stimulation artifacts, neural recording was blanked during stimulation, and for an additional 400 ms, during which the AO remained static.

A dual deep Q network (DQN) reinforcement learning (RL) agent was trained offline in a MATLAB Simulink environment. The mechanical properties (stiffness and damping) of the physical AO-hand system were characterized by advancing the AO through multiple eggshell grasping cycles. These data were curve fitted to determine the relevant properties. A nonlinear Simulink model was then created and used in a simulation environment to train the DQN RL agent.

### Press and Hold

“Press and Hold” assessed the blinded participant’s behavioral and neural responses to cued tactile stimulation of the right wrist and fingers (Fig. 5d). The participant’s right hand was secured to a 45° angled board. Stimuli were applied to four randomized target sites with a force transducer (index pad, index volar pad, thumb pad, Fig. 5c) following a 4s-on, 4s-off paradigm (12 trials per location). The force transducer consisting of a 10 kg miniature tension/compression load cell (DYMH-103, Shanghai Qiyi Co., China) was equipped with a thin rubber tip, a universal inline amplifier (Honeywell International, Inc., USA) for voltage conversion, and the data was recorded using the Noraxon MyoResearch software (Noraxon USA, Inc). The transducer’s output was displayed on the monitor, allowing the experimenter to apply ∼700 g of force. The force sensor was routinely calibrated to maintain accuracy.

The participant reported the perceived location of the stimulus. or reported “Yes” if unsure of the location but felt a sensation, and “No” if no sensation was perceived. Neural data was integrated between 500-5000 Hz to mitigate the effect of speech artefacts in the neural data and were processed as described above.

### S1 cortical mirroring

For each session day, the most recent Q-tip stroking and sensorimotor tasks were used to select the electrodes for stimulation. The neural data was processed as described above and MCCs for each electrode were calculated 0-2s after cue onset in the 100-5000 Hz range for each task, selecting the 16 electrodes with the highest MCC values for each hand/wrist location. Once the Press and Hold experiment started (see Extended Data Fig. 1), electrodes selected through imagination (Q-tip stroking videos) were augmented with electrodes that responded to applied pressure.

Stimulation was applied in intermittent stimulation trains followed by rest periods depending on experimental design and session day. The amplitude, pulse width and interphase stimulation parameters were identical to the S1 mapping paradigm. The frequency used for cortical mirroring (CM) was 200 Hz. Stimulation train times varied between 3.69s-5.67s (5th-95th percentile, max 6s). Rest duration varied between 1.08-3.79s (5th-95th percentile, max 4s). Stimulation trains and rest periods were interleaved. Other tasks targeted elbow flexion and extension (Bicep and Triceps task) and the opening and closing of the hand (Hand Open /Close). For the Bicep and Triceps task the same group of electrodes was stimulated each time. For the Hand Open/Close and Q-Tip task, cue-driven electrode stimulation depending on the condition was applied. Based on MCC values, 10-16 electrodes (minimum MCC > 0.4) were selected for each condition (biceps, triceps, open, close, index tip, thumb tip, wrist), leading to variations in selected electrodes (Extended Data Fig. 5a, b, and c). Performed experiments varied on each session day. Average stimulation time varied between 0 to 25 minutes per electrode per session day (Extended Data Fig. 5d).

### Transcutaneous spinal cord stimulation with cortical mirroring

For the tasks where the participant was receiving both tSCS and cortical mirroring, 10 minutes of tSCS priming over the C5-C6 spinal root was administered followed by a left elbow flexion and extension task (as described above) with tSCS. Next, 5 minutes of combined tSCS and S1 priming (mirrored stimulation patterns performed in S1 without stimuli video or vibrotactile) was applied. Following this, right elbow flexion and extension task was performed with continuous tSCS, while cued S1-ICMS was applied only during the active flexion/extension cues (not during rest or inter-set rest). Both tSCS and S1-ICMS were discontinued after the final right-side set. No cue-driven S1-ICMS was applied during the left-side sets.

### S1 cortical mirroring with vibrotactile stimulation

When the participant received both cortical mirroring and vibrotactile stimulation, depending on the cue, electrodes of either index tip, thumb tip or wrist were stimulated. Simultaneously, vibrotactile stimulation was applied to the same locations and applied using a battery-powered ESP32 microcontroller and four haptic motor controllers (DRV2605L, Adafruit Industries) each connected to a mini vibrating motor disc (Model 10B27.3018, 100614; 10 mm diameter x 2.7 mm thickness, Adafruit Industries).

## Data Availability

All data produced in the present study are available upon reasonable request to the authors

## Acknowledgements

The authors wish to express their sincere gratitude to the participant of this study, along with his family and aides for their invaluable contributions. We also extend our thanks to the clinical and administrative staff of Northwell Health and specifically Dan Sciubba and the Northwell Neurosurgery, and North Shore University Hospital for their support and excellence in preparation for and during the surgery, the Northwell Health Radiology Department for their help on imaging, and the Institute for Bioelectronic Medicine administrative staff for their administrative support. We also acknowledge the contributions of everyone involved in the custom active orthosis 3D printing process, Nikunj Bhagat, Sadegh Ebrahimi, Jose Herrero-Rubio, Elizabeth Espinal, Noah Markowitz, Xiang He, Todd Lefkowitz, Emma Ronai, and Dana Fried for their assistance during the study. We would also like to thank Rob Franklin and Greg Palis, both from Blackrock Neurotech for their expertise and technical guidance throughout the study.

## Author contributions

C.E.B., S.C., and R.R. conceptualized the study.

S.C. and C.E.B. designed the experiments and led the investigation.

E.I. and R.R. managed regulatory affairs and participant session logistics.

E.I., R.R., D.G., and C.M. performed clinical assessments.

S.C., S.K.W., E.I., I.A.R. collected data and performed formal analysis

A.J. and Z.E. led the hardware and software engineering.

A.J., Z.E., A.N., T.A.G., and C.E.B led the development of the active orthosis.

S.C., S.K.W., A.J., optimized the software and real-time pipeline

S.C. and R.R. led the fMRI scans. J-W.K and J.X. analyzed the fMRI data.

S.C., C.E.B, with the assistance of M.F.G., planned the initial array locations.

A.D.M., N.B-S., S.C., C.E.B., N.E.C., M.S.F., M.F., and N.G.C performed pre-surgical planning.

G.T. and S.B. imported the planned array locations into the surgical navigation system.

A.D.M., N.B-S. and R.P. performed the surgery for array implantation, with M.F. and N.G.C. on anesthesia.

G.T., S.B. and A.D.M. performed intraoperative mapping.

C.E.B., A.B.S., and A.D.M. supervised and guided the overall study.

S.C., S.K.W., A.J., Z.E., E.I., I.A.R., and C.E.B. wrote the original draft.

All authors reviewed and edited the manuscript.

## Ethics Declaration/Competing Interests

CEB has financial interests in Neuvotion, Inc, a medical device company developing neurotechnologies for restoring function after stroke or spinal cord injury, and Sanguistat, LLC, a company developing neurostimulation technologies to reduce blood loss; he also has multiple patents in the field of neuroprosthetics and related fields. The remaining authors declare no competing interests.

## Extended Data Figures and Tables

**Extended Data Fig. 1:**
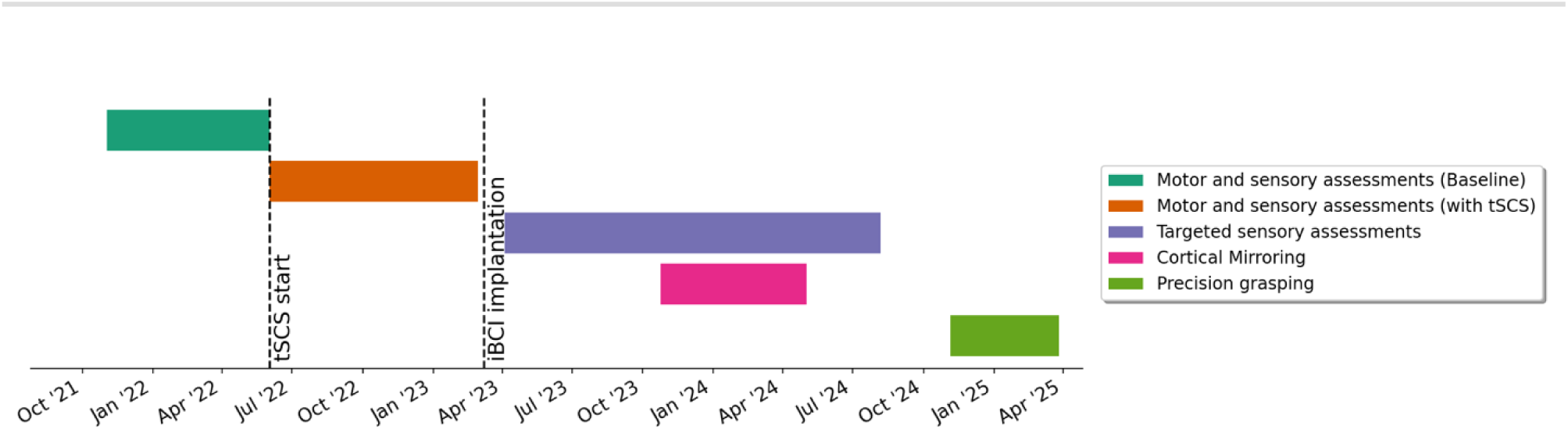
Study timeline. This shows the different phases of the study presented in this work and the timelines of data acquisition for each of the study experiments. The participant sustained a SCI 13 months prior to the start of the study.

**Extended Data Fig. 2:**
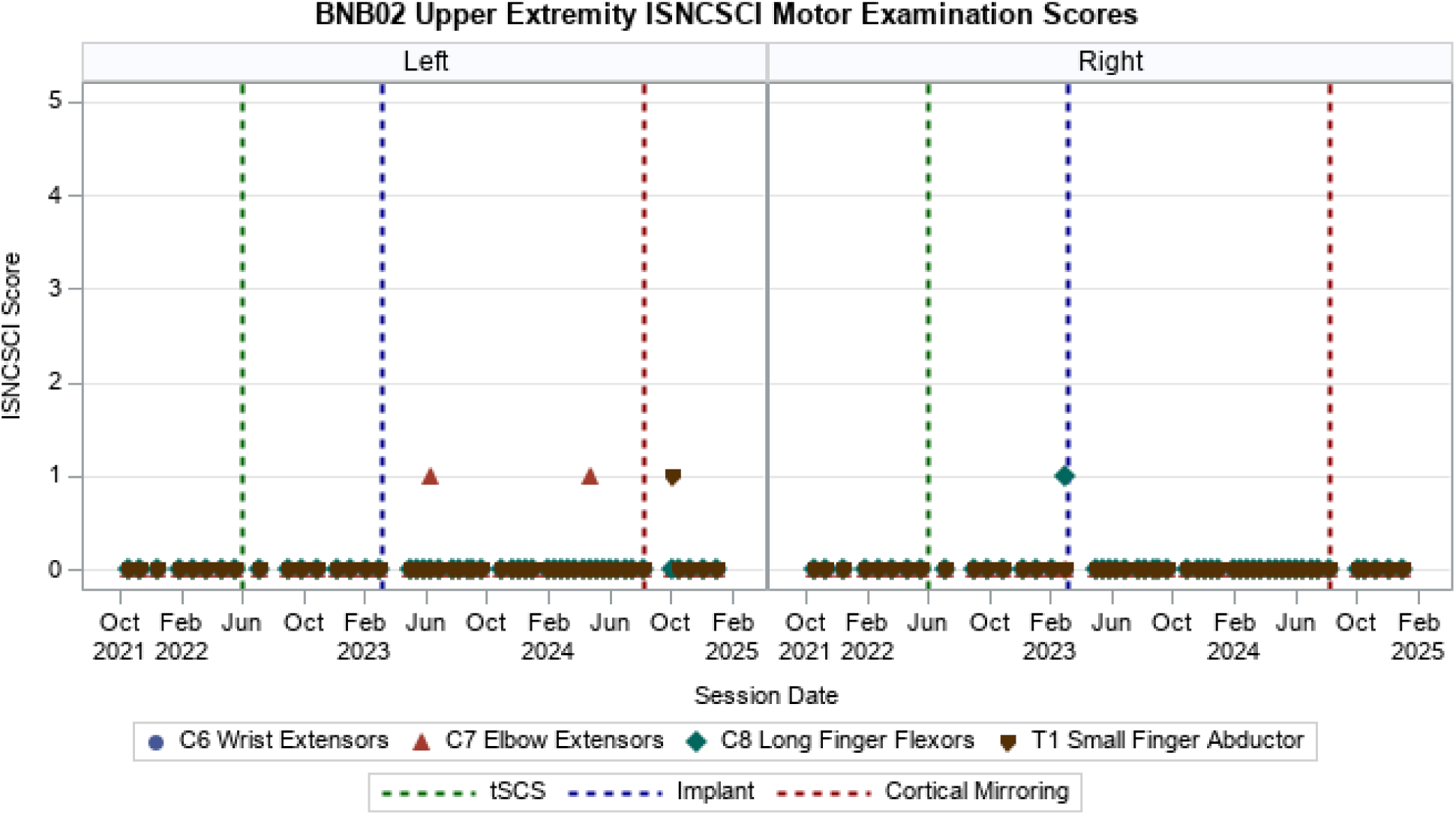
Upper extremity ISNCSCI motor examination scores. Scoring criteria: 0 = Total paralysis; 1 = Palpable or visible contraction; 2 = Active movement, full range of motion (ROM) with gravity eliminated; 3 = Active movement, full ROM against gravity; 4 = Active movement, full ROM against gravity and moderate resistance in a muscle specific position; 5 = (Normal) active movement, full ROM against gravity and full resistance in a functional muscle position expected from an otherwise unimpaired person. ISNCSCI = International Standards for Neurological Classification of Spinal Cord Injury

**Extended Data Fig. 3:**
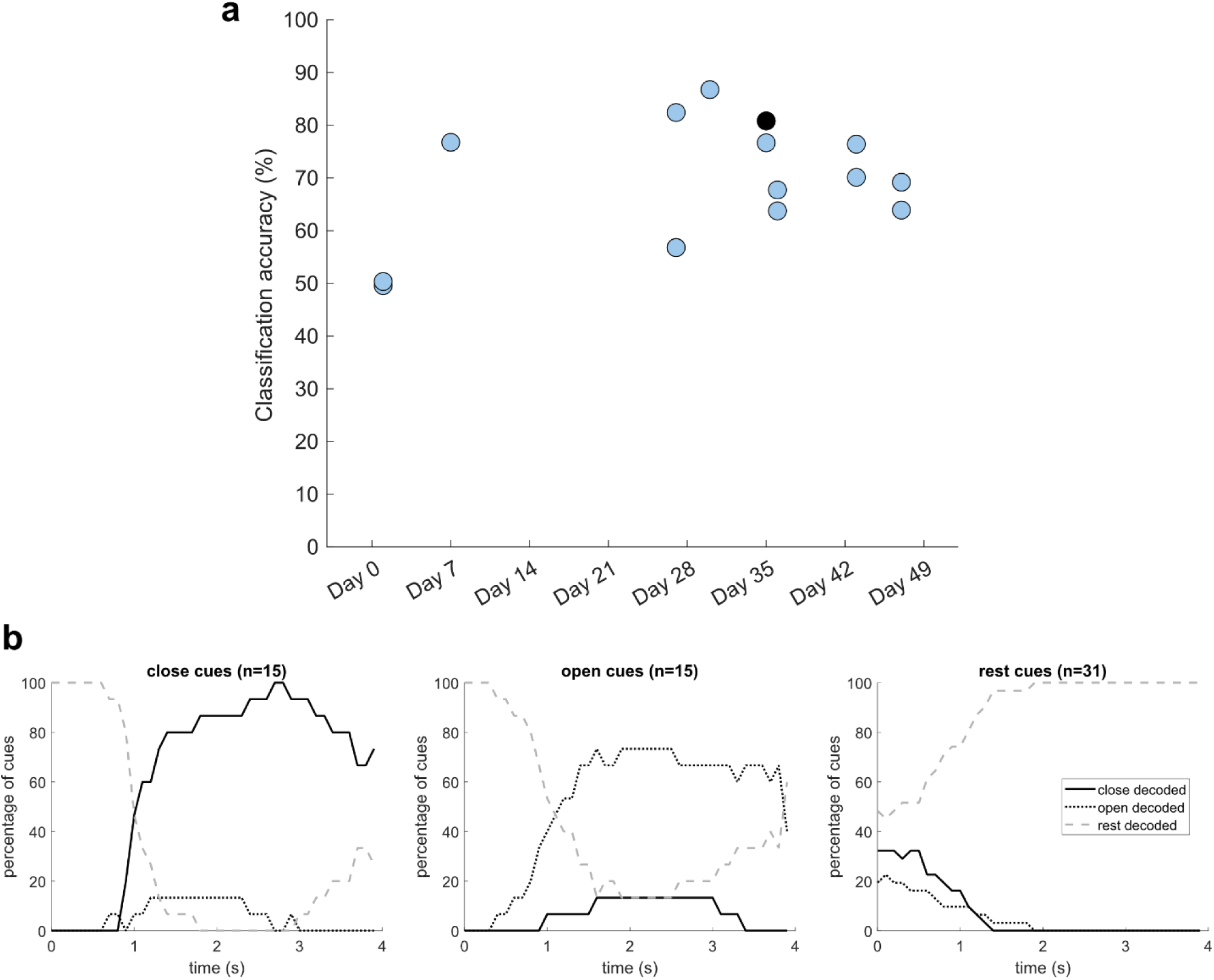
Decoding of attempted hand open and close (see Fig. 3b). **a)** Accuracies for decoding of hand state (close, open, rest) over 15 data blocks collected over 8 session days spanning more than a month of time. Chance = 33.3%. Example data block shown in (b) shown in black. **b)** Example decoding performance for one experimental block (overall accuracy=80.8%). For each cue type, over 4 seconds from the start of that cue, the percentage of trials in each decoder state is shown. The decoder performance could be used to trigger NMES stimulation.

**Extended Data Fig. 4:**
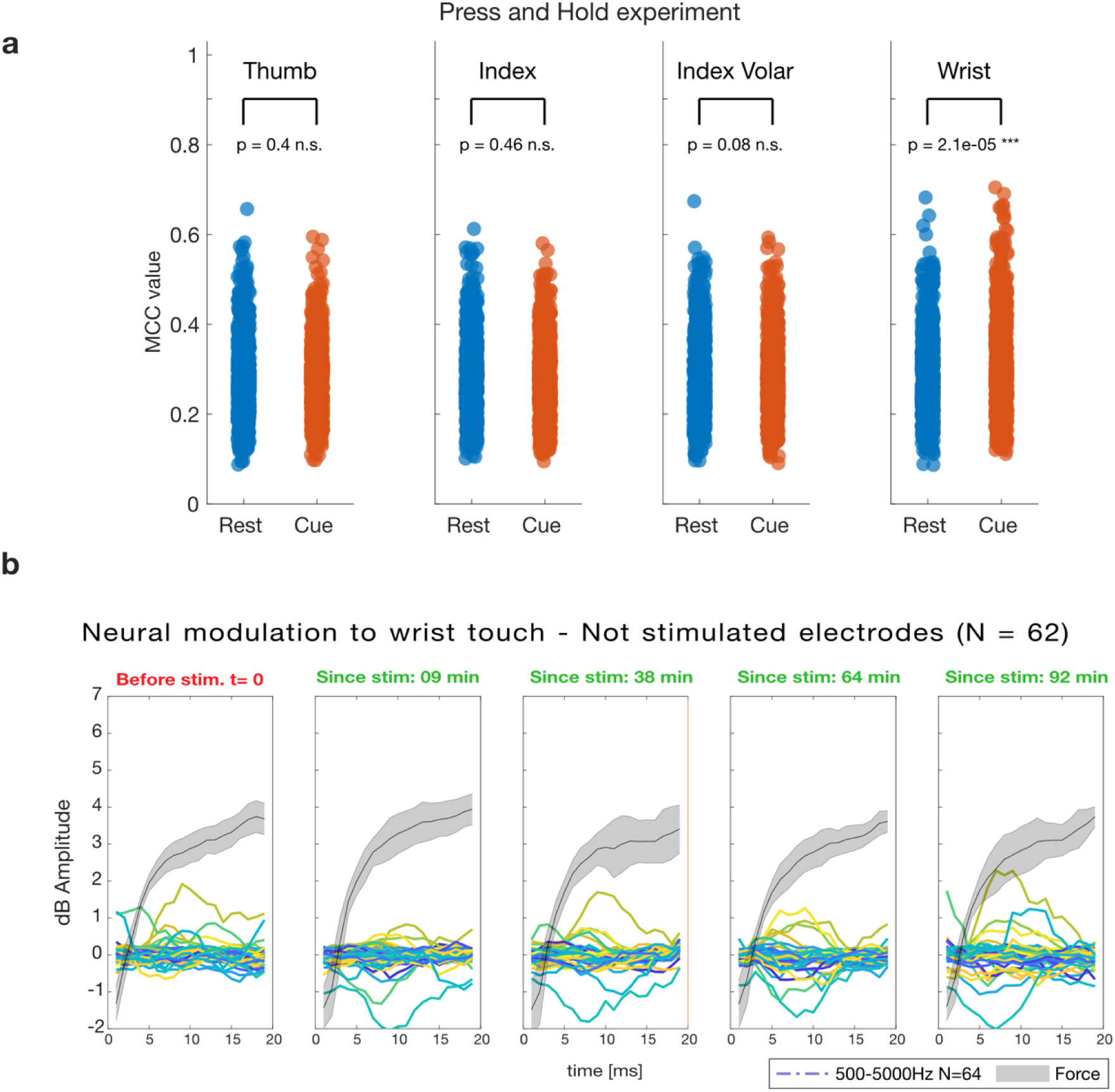
Neural modulation to Press and Hold task. **a)** MCC comparison of 960 electrodes (96 x 10 sessions) during a 1.4s period during rest vs. a 1.4s period during applied pressure to the thumb pad, index pad, index volar pad and wrist. Only the wrist demonstrated a significant difference in activity (paired t-test, p < 0.001). **b)** Effect of time since stimulation on feature amplitude in not stimulated electrodes (N = 64, one color per electrode). Neural data was aligned to force onset for each trial (mean and std. of force). Average electrode activity over all trials was plotted for 0.2 s before and 1.6s after force onset.

**Extended Data Fig. 5:**
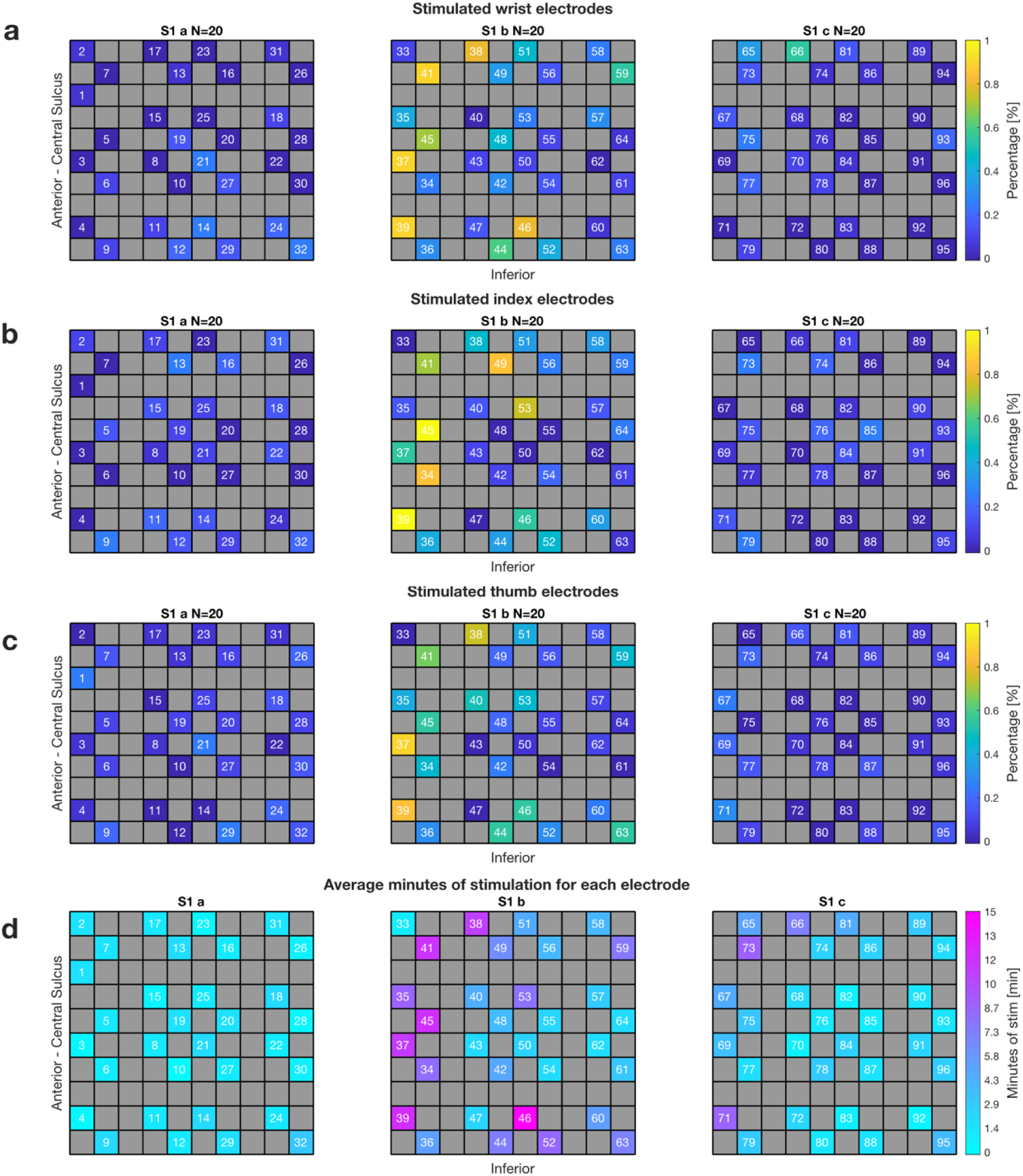
Selected electrodes for cortical mirroring. **a-c)** Percentage of selection for stimulation for each S1 at the (**a**) wrist, (**b**) index pad and (**c**) thumb pad location. **d)** Average stimulation time (in minutes) per session for each electrode, reflecting total stimulation throughout the entire session day, not limited to hand location.

## Supplementary Information

Nature_SuppVids_Ver2_NMES.mp4

**Supplementary Video 1.** Participant demonstrates cortically mediated light grasping through neuromuscular electrical stimulation. Available upon request

Nature_SuppVids_Ver2_AO.mp4

**Supplementary Video 2.** Participant demonstrates cortically mediated precision grasping and self-feeding using an active orthosis. Available upon request

**Supplementary Table 1.**
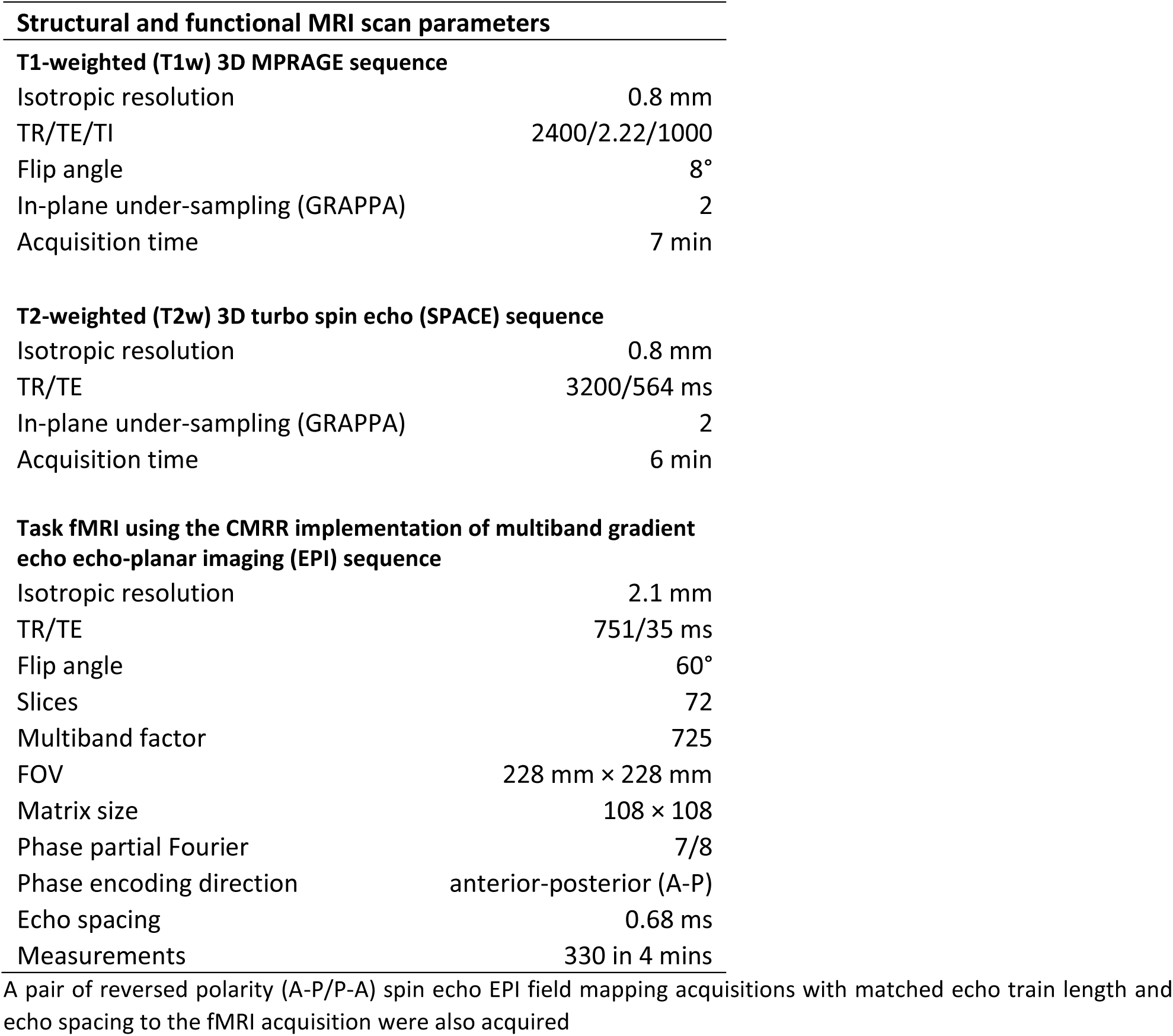
Optimized structural and functional MRI scan parameters for localizing hand regions in the primary motor and somatosensory cortices.

## References

1. University of Alabama at Birmingham. 2023 Annual Statistical Report for the Spinal Cord Injury Model Systems. (2023).

2. Armour, B. S., Courtney-Long, E. A., Fox, M. H., Fredine, H. & Cahill, A. Prevalence and Causes of Paralysis—United States, 2013. Am J Public Health 106, 1855–1857 (2016).

3. Kirshblum, S., Snider, B., Eren, F. & Guest, J. Characterizing Natural Recovery after Traumatic Spinal Cord Injury. Journal of Neurotrauma 38, 1267–1284 (2021).

4. Bouton, C. E. et al. Restoring cortical control of functional movement in a human with quadriplegia. Nature 533, 247–250 (2016).

5. Ajiboye, A. B. et al. Restoration of reaching and grasping in a person with tetraplegia through brain-controlled muscle stimulation: a proof-of-concept demonstration. Lancet 389, 1821–1830 (2017).

6. Flesher, S. N. et al. Intracortical microstimulation of human somatosensory cortex. Sci Transl Med 8, 361ra141 (2016).

7. Osborn, L. E. et al. Subthreshold intracortical microstimulation of human somatosensory cortex enhances tactile sensitivity. medRxiv 2024.06.21.24309202 (2024) doi:10.1101/2024.06.21.24309202.

8. Lorach, H. et al. Walking naturally after spinal cord injury using a brain–spine interface. Nature 618, 126–133 (2023).

9. Glasser, M. F. et al. A multi-modal parcellation of human cerebral cortex. Nature 536, 171–178 (2016).

10. Chandrasekaran, S. et al. Targeted transcutaneous spinal cord stimulation promotes persistent recovery of upper limb strength and tactile sensation in spinal cord injury: a pilot study. Front. Neurosci. 17, (2023).

11. Bouton, C. et al. Decoding Neural Activity in Sulcal and White Matter Areas of the Brain to Accurately Predict Individual Finger Movement and Tactile Stimuli of the Human Hand. Frontiers in Neuroscience 15, (2021).

12. Oya, T., Takei, T. & Seki, K. Distinct sensorimotor feedback loops for dynamic and static control of primate precision grip. Commun Biol 3, 156 (2020).

13. Anderson, K. D. Targeting Recovery: Priorities of the Spinal Cord-Injured Population. Journal of Neurotrauma 21, 1371–1383 (2004).

14. Flesher, S. N. et al. A brain-computer interface that evokes tactile sensations improves robotic arm control. Science 372, 831–836 (2021).

15. Lin, A. et al. A Review of Functional Restoration From Spinal Cord Stimulation in Patients With Spinal Cord Injury. Neurospine 19, 703–734 (2022).

16. Chandrasekaran, S. et al. Case study: persistent recovery of hand movement and tactile sensation in peripheral nerve injury using targeted transcutaneous spinal cord stimulation. Frontiers in Neuroscience 17, (2023).

17. Moritz, C. et al. Non-invasive spinal cord electrical stimulation for arm and hand function in chronic tetraplegia: a safety and efficacy trial. Nat Med 30, 1276–1283 (2024).

18. Huang, R. et al. Minimal handgrip force is needed for transcutaneous electrical stimulation to improve hand functions of patients with severe spinal cord injury. Scientific Reports 2022 12:1 12, 1–13 (2022).

19. Asan, A. S., McIntosh, J. R. & Carmel, J. B. Targeting Sensory and Motor Integration for Recovery of Movement After CNS Injury. Front. Neurosci. 15, (2022).

20. Rosenthal, I. A. et al. S1 represents multisensory contexts and somatotopic locations within and outside the bounds of the cortical homunculus. Cell Reports 42, 112312 (2023).

21. Bashford, L. et al. The Neurophysiological Representation of Imagined Somatosensory Percepts in Human Cortex. J. Neurosci. 41, 2177–2185 (2021).

22. Song, W., Kerr, C. C., Lytton, W. W. & Francis, J. T. Cortical Plasticity Induced by Spike-Triggered Microstimulation in Primate Somatosensory Cortex. PLOS ONE 8, e57453 (2013).

23. Rosén, B. & Lundborg, G. A model instrument for the documentation of outcome after nerve repair. J Hand Surg Am 25, 535–543 (2000).

24. Glasser, M. F. et al. The Minimal Preprocessing Pipelines for the Human Connectome Project. Neuroimage 80, 105–124 (2013).

25. Robinson, E. C. et al. MSM: A new flexible framework for Multimodal Surface Matching. NeuroImage 100, 414–426 (2014).

26. Robinson, E. C. et al. Multimodal surface matching with higher-order smoothness constraints. NeuroImage 167, 453–465 (2018).

27. Griffanti, L. et al. ICA-based artefact removal and accelerated fMRI acquisition for improved resting state network imaging. NeuroImage 95, 232–247 (2014).

28. Salimi-Khorshidi, G. et al. Automatic denoising of functional MRI data: Combining independent component analysis and hierarchical fusion of classifiers. NeuroImage 90, 449–468 (2014).

29. Glasser, M. F. et al. Using temporal ICA to selectively remove global noise while preserving global signal in functional MRI data. NeuroImage 181, 692–717 (2018).

30. Woolrich, M. W., Ripley, B. D., Brady, M. & Smith, S. M. Temporal Autocorrelation in Univariate Linear Modeling of FMRI Data. NeuroImage 14, 1370–1386 (2001).

31. Nanivadekar, A., Chandrasekaran, S., Gaunt, R. & Fisher, L. RNEL PerceptMapper. Zenodo (2020) 10.5281/zenodo.3939658.

